# Implementation and Evaluation of a Home-Based Pre-Exposure Prophylaxis Monitoring Option: A Protocol for a Randomized Controlled Trial

**DOI:** 10.1101/2023.12.17.23300112

**Authors:** Chase A. Cannon, Kate Holzhauer, Matthew Golden

## Abstract

**Background:** HIV prevention is a public health priority. Despite progress in recent years, PrEP use remains suboptimal especially among groups disproportionately impacted by new HIV diagnoses such as gender and sexual minorities of color. Multiple barriers including lack of PrEP providers and challenges with attending quarterly monitoring visits contribute to low PrEP uptake and retention. Home-based PrEP (HB-PrEP) services could reduce stigma, increase convenience, expand health system capacity for PrEP care, and improve PrEP retention.

**Objective:** HOT4PrEP (Home Option Testing for PrEP) is a hybrid randomized controlled trial (RCT) that aims to examine whether HB-PrEP care is acceptable to PrEP users, feasible to implement into a sexual health clinic setting, and impacts PrEP retention over time.

**Methods:** The RCT will recruit approximately 450 persons currently taking or soon to initiate PrEP at the Sexual Health Clinic in Seattle, WA and randomize them to continue standard of care or have the option to use HB-PrEP for two of three tri-annual PrEP follow-up visits. Participants in the intervention arm will be sent home kits containing gonorrhea and chlamydia swabs and Tasso devices for blood self-collection. The primary outcome is PrEP retention between groups at 20 months; secondary outcomes include user satisfaction/acceptability, feasibility, self-reported PrEP adherence, and STI incidence. Interviews with PrEP users and clinic staff will elucidate barriers and facilitators of implementation.

**Results:** The HOT4PrEP RCT began enrolling in March 2022, was on hold during the height of the US mpox epidemic, then resumed in December 2022. From the first 100 enrollees, median age is 34 years and most are cisgender gay men (89%) with at least some college education (91%). Among those randomized to the HB-PrEP option, 65% have opted at least once to self-collect samples at home and 84% have successfully returned test kits for HIV/STI testing. Primary PrEP retention and qualitative analyses are ongoing.

**Conclusions:** Implementation of HB-PrEP into a high-volume sexual health clinic seems to be feasible and acceptable to early RCT enrollees. This strategy has the potential to address individual and systemic barriers associated with initiating and persisting on PrEP such as increasing sexual health agency and expanding clinical capacity to serve greater numbers of PrEP users.

Trial Registration: ClinicalTrials.gov NCT05856942

## Introduction

### Background

In 2021, the majority of new human immunodeficiency virus (HIV) infections in the United States (US) occurred in sexual minority men (SMM).(1) The US federal government targeted resources toward the goal of reducing new HIV infections by 75% by the year 2025 through the “Ending the HIV Epidemic” (EHE) initiative(2) and highlights prevention of new infections as a key component of the plan. While pre-exposure prophylaxis (PrEP) has been proven to reduce new HIV diagnoses at the population level,(3,4) it remains an underutilized resource. Preliminary evidence suggests that only 36% of people in the US with an indication for PrEP received a prescription in in 2022,(5) up only slightly from 30% in 2021.(6) Though PrEP coverage among SMM at greatest risk for HIV in Seattle-King County, Washington (WA) approached 65% in 2021,(7) PrEP use remains low among younger people and some SMM of color.(4,7) As part of its EHE initiatives, Public Health-Seattle & King County (PHSKC) aims to achieve ≥70% PrEP coverage among SMM and transgender persons, with equal use among all racial/ethnic groups.

Structural barriers such as lack of access to PrEP medical providers and the burden of quarterly clinical monitoring visits have stymied PrEP scale-up efforts in the US for years.(8–10) PrEP retention (defined as maintaining all aspects of PrEP care, including attending follow-up visits and obtaining monitoring tests every 3 months according to national guidelines(11)) represents a substantial gap in the PrEP care continuum.(12,13) Studies on national PrEP retention rates suggest only 56% of individuals remain on PrEP from initiation to year 1 and only 41% persist through year 2.(14) Younger age, Black race and unstable or lower income are each associated with discontinuations and being lost to follow-up.(15) We have observed similar trends at PHSKC’s Sexual Health Clinic (SHC) in that 40% of PrEP users discontinue at least once at 12 months.(16) The COVID-19 pandemic created dramatic reductions in sexual health service capacity(17) that only further exacerbated and continues to impact existing structural barriers that lead to lower PrEP access and higher numbers of discontinuations.(18–20) Among SMM newly diagnosed with HIV in King County, prior PrEP use and discontinuations are common and underscore the urgent need to develop and implement novel targeted strategies to minimize barriers and provide individuals with more options to stay on PrEP.(21)

Home-based PrEP (HB-PrEP) – whereby an individual collects PrEP monitoring specimens at home, mails them into an affiliated laboratory for testing, and follows up with a provider remotely to receive a renewed PrEP prescription – has the potential to decrease PrEP access barriers and increase PrEP continuation rates. Studies confirm that patients are comfortable self-collecting bacterial sexually transmitted infection (STI) screening specimens(22) and that HB-PrEP monitoring is acceptable and often preferable to in-person care.(12,23,24) Configuring PrEP programs to users’ needs can optimize both uptake and delivery.(25) Particularly for young SMM, programs that are convenient, discreet, and provide them with agency over PrEP use and monitoring may reduce the stigma associated with PrEP use, and in turn, increase their likelihood of PrEP uptake and retention.(12,24) Moreover, HB-PrEP may decongest high-volume clinical sites and create additional capacity to grow PrEP programs.

Several direct-to-consumer companies(26) and fewer research or clinic-based PrEP programs provide PrEP services at home.(23,24,27) However, opportunities to adequately screen for newly acquired syphilis and HIV in the home setting are often limited. Given that STI incidence is high among people on PrEP,(28) US Centers for Disease Control & Prevention (CDC) guidelines recommend PrEP users be screened for STI quarterly, and specifically for syphilis and HIV using assays with narrow windows of detection.(13) Syphilis is highly associated with HIV acquisition in PrEP users;(29) thus, it is important that PrEP monitoring include tests that can distinguish new from old syphilis infections, such as with the quantitative rapid plasma reagin (RPR). Approximately 40% of PHSKC PrEP users have a history of syphilis, of which 25% are serofast (have a persistently positive RPR despite treatment in the past) and require a quantitative RPR for syphilis screening.(30) Although prior HB-PrEP studies used procedures that allowed PrEP users to collect all specimens (gonorrhea/chlamydia [GC/CT] swabs and blood samples) from home,(23,27) nearly all require fingerstick specimens to perform a less accurate rapid HIV antibody test and collected insufficient blood volume to perform RPR titers, limiting the ability to determine whether titers were stable or signified a newly acquired syphilis infection. We previously conducted a pilot study(30) that showed the acceptability and feasibility of using a novel blood self-collection device (Tasso+™)(31) to obtain samples suitable for HIV antigen/antibody and quantitative RPR testing. However, whether these devices could be used successfully within a dedicated program for serial PrEP monitoring remains unknown.

### Objectives

Through a hybrid(32) randomized controlled trial (RCT), we aim to simultaneously evaluate the effectiveness of HB-PrEP as a strategy for improving PrEP retention and its implementation into an urban SHC’s existing PrEP program.

The specific aims of our study are to:

∘ Aim 1: Evaluate the impact of HB-PrEP on PrEP retention rates over time among groups assigned to either home-based monitoring or routine care;
∘ Aim 2: Assess the program’s reach and define the factors that influence HB-PrEP implementation.

The following study procedures have been reviewed and approved by the University of Washington Institutional Review Board (STUDY00013871) and were considered not to pose any more than minimal risk to study participants.

## Methods

### Study Overview

The HOT4PrEP (Home Option Testing for PrEP) project is a series of interrelated studies designed to implement and evaluate a HB-PrEP monitoring system that includes the option for biological specimen self-collection at home and remote clinical follow-up in Seattle-King County, WA. A detailed description of Tasso device use and results of pre-implementation acceptability and feasibility analyses are published elsewhere.(30) The current phase of the HOT4PrEP project consists of two components: (1) a hybrid RCT wherein we evaluate HB-PrEP as an implementation strategy and its effect on PrEP retention over time along with other secondary outcomes, and (2) a mixed-methods analysis to assess reach and both individual and system-level barriers & facilitators (B&F) to HB-PrEP implementation.

For the first study component, we will enroll 458 participants from the PHSKC SHC in a 1:1 randomized fashion to either continue PrEP standard of care [(SOC), control group] or to have a HB-PrEP monitoring option (intervention). Participants in both arms will complete serial online surveys and receive PrEP follow-up via telehealth modalities or at the clinic (Table 1). At the time of trial conception, monitoring intervals were initially set at every 3 months; however, these changed to every 4 months in March 2023 following the SHC PrEP program’s shift to reduce the total number of required annual visits. HB-PrEP participants receive mailed sampling kits containing instructions and materials to collect extragenital (pharyngeal and rectal) GC/CT swabs and two devices to self-collect capillary blood from the upper arm into microtainer tubes. Kits are returned via postal mail to the PHSKC laboratory for creatinine, quantitative syphilis serologies and HIV antigen/antibody testing. Secondary outcomes include reach (extent of kit uptake and participation in the program), user satisfaction with the assigned PrEP monitoring program, STI incidence, and time from resulting of abnormal laboratory values to participant notification. Participation in this first RCT component is not incentivized.

**Table 1:**
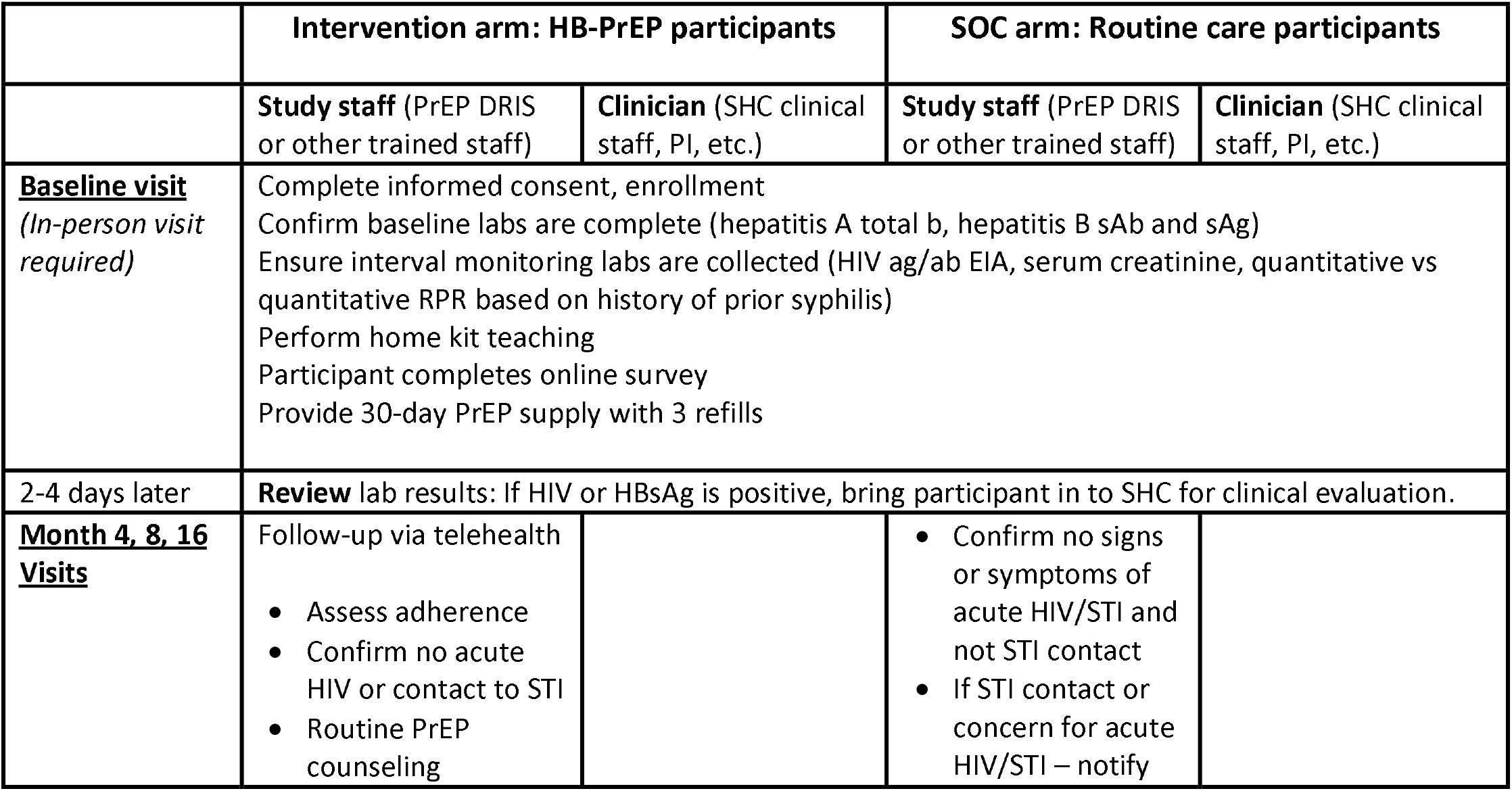
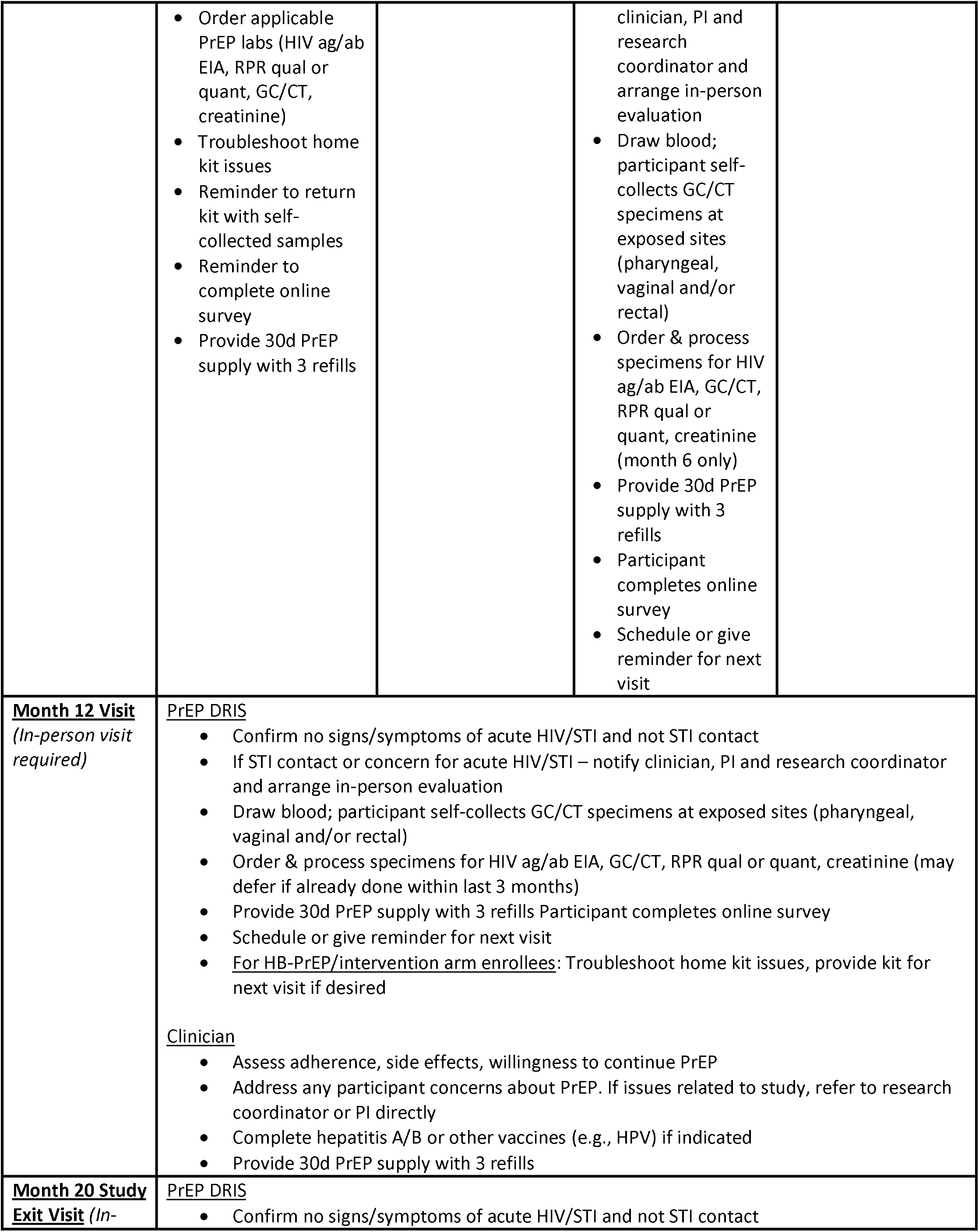
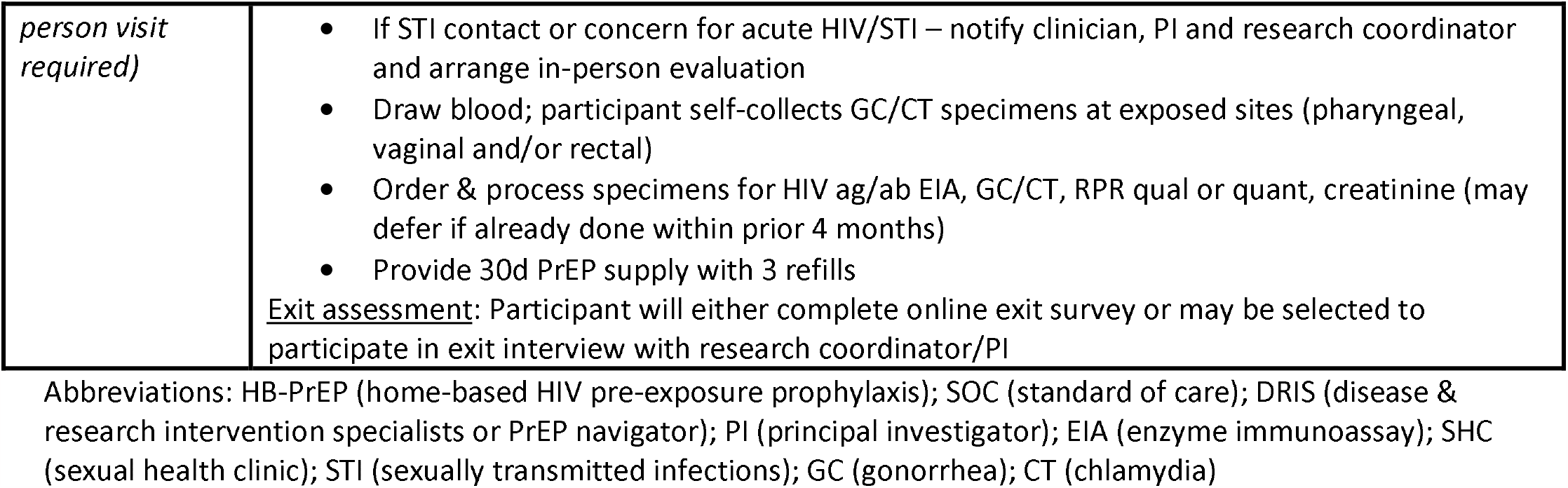
HOT4PrEP RCT Study Procedures.

|  | Intervention arm: HB-PrEP participants |  | SOC arm: Routine care participants |  |
| --- | --- | --- | --- | --- |
|  | Study staff (PrEP DRIS or other trained staff) | Clinician (SHC clinical staff, PI, etc.) | Study staff (PrEP DRIS or other trained staff) | Clinician (SHC clinical staff, PI, etc.) |
| **<u>Baseline visit</u>**<br>(In-person visit required) | Complete informed consent, enrollment<br>Confirm baseline labs are complete (hepatitis A total b, hepatitis B sAb and sAg)<br>Ensure interval monitoring labs are collected (HIV ag/ab EIA, serum creatinine, quantitative vs quantitative RPR based on history of prior syphilis)<br>Perform home kit teaching<br>Participant completes online survey<br>Provide 30-day PrEP supply with 3 refills |  |  |  |
| 2-4 days later | **Review** lab results: If HIV or HBsAg is positive, bring participant in to SHC for clinical evaluation. |  |  |  |
| **<u>Month 4, 8, 16 Visits</u>** | Follow-up via telehealth<br>Assess adherence<br>Confirm no acute HIV or contact to STI<br>Routine PrEP counseling |  | Confirm no signs or symptoms of acute HIV/STI and not STI contact<br>If STI contact or concern for acute HIV/STI – notify |  |
|  | • Order applicable PrEP labs (HIV ag/ab EIA, RPR qual or quant, GC/CT, creatinine)<br>• Troubleshoot home kit issues<br>• Reminder to return kit with self-collected samples<br>• Reminder to complete online survey<br>• Provide 30d PrEP supply with 3 refills |  | clinician, PI and research coordinator and arrange in-person evaluation<br>• Draw blood; participant self-collects GC/CT specimens at exposed sites (pharyngeal, vaginal and/or rectal)<br>• Order & process specimens for HIV ag/ab EIA, GC/CT, RPR qual or quant, creatinine (month 6 only)<br>• Provide 30d PrEP supply with 3 refills<br>• Participant completes online survey<br>• Schedule or give reminder for next visit |  |
| **Month 12 Visit**<br>*(In-person visit required)* | <u>PrEP DRIS</u><br>• Confirm no signs/symptoms of acute HIV/STI and not STI contact<br>• If STI contact or concern for acute HIV/STI – notify clinician, PI and research coordinator and arrange in-person evaluation<br>• Draw blood; participant self-collects GC/CT specimens at exposed sites (pharyngeal, vaginal and/or rectal)<br>• Order & process specimens for HIV ag/ab EIA, GC/CT, RPR qual or quant, creatinine (may defer if already done within last 3 months)<br>• Provide 30d PrEP supply with 3 refills Participant completes online survey<br>• Schedule or give reminder for next visit<br>• <u>For HB-PrEP/intervention arm enrollees</u>: Troubleshoot home kit issues, provide kit for next visit if desired<br><u>Clinician</u><br>• Assess adherence, side effects, willingness to continue PrEP<br>• Address any participant concerns about PrEP. If issues related to study, refer to research coordinator or PI directly<br>• Complete hepatitis A/B or other vaccines (e.g., HPV) if indicated<br>• Provide 30d PrEP supply with 3 refills |  |  |  |
| **Month 20 Study Exit Visit** *(In-* | <u>PrEP DRIS</u><br>• Confirm no signs/symptoms of acute HIV/STI and not STI contact |  |  |  |

|  |  |
| --- | --- |
| *person visit required)* | • If STI contact or concern for acute HIV/STI – notify clinician, PI and research coordinator and arrange in-person evaluation<br>• Draw blood; participant self-collects GC/CT specimens at exposed sites (pharyngeal, vaginal and/or rectal)<br>• Order & process specimens for HIV ag/ab EIA, GC/CT, RPR qual or quant, creatinine (may defer if already done within prior 4 months)<br>• Provide 30d PrEP supply with 3 refills<br><u>Exit assessment:</u> Participant will either complete online exit survey or may be selected to participate in exit interview with research coordinator/PI |

Abbreviations: HB-PrEP (home-based HIV pre-exposure prophylaxis); SOC (standard of care); DRIS (disease & research intervention specialists or PrEP navigator); PI (principal investigator); EIA (enzyme immunoassay); SHC (sexual health clinic); STI (sexually transmitted infections); GC (gonorrhea); CT (chlamydia)

For the second component of the study, we will conduct a series of in-depth interviews with study participants, staff and administrative personnel to determine factors associated with the program’s implementation. Participation in interviews for this second study component will be compensated.

### Study Setting and Staff

The PHSKC SHC provides direct medical services related to sexual health and STI to over 6,500 patients each year and is the largest single diagnosing site for HIV infection in the Pacific Northwest region of the US. Of all patients seen annually in the SHC, approximately 41% identify as SMM and 42% as racial/ethnic minorities. Disease & Research Intervention Specialists (DRIS), who are non-clinical public health staff specially trained in HIV/STI prevention and to provide partner services, currently serve as PrEP navigators and conduct all PrEP initiation and non-clinical monitoring visits in the SHC.(33) They will continue to fulfill this role during the study, augmenting their current work to manage HB-PrEP, and along with the lead research coordinator will help with eligibility screening, recruitment and retention efforts.

### Primary RCT

#### Eligibility Criteria and Exclusions

All patients in the SHC meet both PrEP use criteria according to CDC’s 2021 clinical practice guidelines (13) and at least one of three PHSKC PrEP criteria: (1) HIV-negative cisgender man, transgender or nonbinary person who has sex with men; (2) person who has a partner with unsuppressed HIV; or (3) person who injects drugs or engages in exchange sex.(33) Prospective participants for the trial additionally must meet the following study-specific criteria.

### Inclusion criteria

(1) Age ≥18 years; (2) resident of Washington State; (3) can speak, understand, read and/or write in English or Spanish; (4) current PrEP user or interested in starting or restarting PrEP; (5) documented negative HIV antigen/antibody test within the last 4 weeks (or confirmation of consistent self-reported PrEP use since last HIV ag/ab test within prior 3 months); (6) willingness to be randomized to HB-PrEP and adhere to specified procedures; (7) willingness to provide primary and alternate contact information.

### Exclusion criteria

(1) Recent (<4 weeks) “high risk” HIV exposure while off PrEP or symptoms of acute HIV infection; (2) no mailing address to receive packages or sampling kits; (3) no stable, working telephone number; (4) no smartphone or electronic device with internet access; (5) history of bleeding disorder, current or recent (≤7 days) use of anticoagulant medications (warfarin, rivaroxaban, etc.); (6) pregnancy.

Persons with active hepatitis B or a creatinine clearance <50 mL/min are not excluded. However, they will receive counseling about the risk of hepatitis B infection worsening if PrEP medications are discontinued and be encouraged to follow up with medical providers outside of the study for management of comorbid conditions.

#### Screening, Recruitment and Enrollment

##### Existing patients

DRIS and/or the research staff review patients scheduled for routine PrEP visits at the SHC and offer enrollment in person to all eligible patients. Enrollees are randomized to continue SOC (attend the PHSKC SHC every 4 months for a visit with DRIS, have blood samples collected for PrEP monitoring, and perform self-collection of gonorrhea/chlamydia swabs) or have the option to do HB-PrEP monitoring. Those assigned to HB-PrEP (intervention arm) are eligible to participate in home-based monitoring immediately after receiving instruction from study staff on how to use the Tasso devices, procedures for mailing kits back to the PHSKC laboratory, and how to complete online surveys after returning kits.

##### New and former (“lost to follow up”) PHSKC PrEP patients

Prospective and former PrEP patients who are eligible for the study may be recruited at their initial visit. Those randomized to the control arm will adhere to usual SHC PrEP clinical protocols.(33) Persons randomized to HB-PrEP receive instruction from DRIS or study staff about self-collection procedures as above and are eligible to return their first kit for the month 4 monitoring cycle. Persons initiating PrEP (or re-initiating after a prolonged discontinuation) must be evaluated in person by a clinician and have blood collected by clinic staff for HIV screen, renal function, viral hepatitis serologies, etc.; thus, these individuals will become eligible to use home kits at their first tri-annual follow-up (i.e., 120 days after PrEP initiation). Person-time in the study for all subjects begins immediately after enrollment.

#### Randomization and Study Procedures

##### Sample size and power considerations

Assuming that 2.5% of individuals discontinue PrEP per month in the control arm, we expect 45% of SOC subjects to discontinue PrEP (55% retention) over the 20-month study period. In the intervention arm we estimate 70% will adopt HB-PrEP. We assume a relative risk of discontinuation of 0.6 among adopters leading to a 32% discontinuation rate (68% retention) in the HB-PrEP arm. We also allowed for an overall dropout rate in both groups of 6%.(16) Thus, a sample size of 458 participants should yield 80% power to detect a 13%-point difference in PrEP retention. All eligible participants are assigned via simple 1:1 randomization to either the intervention arm (n=229) or SOC arm (n=229) and followed for 20 months. RCT procedures are detailed in Table 1.

##### Surveys

Participants in both arms are asked to complete online surveys using REDCap (Research Electronic Data Capture)(34)—a secure, web-based platform that participants used successfully in the pre-implementation pilot study. The initial enrollment survey collects basic demographic and contact information including valid mailing address and phone number for the participant and an approved alternative contact, sexual orientation and gender identity items, and PrEP monitoring preferences. Subsequent interval surveys focus on evaluating self-reported PrEP adherence, number of sex partners, type and frequency of sex, frequency of drug or condom use, and measures of satisfaction with the assigned monitoring program.

##### Home specimen tracking

Home sampling kits contain instruction sheets in English or Spanish, specimen labels, two swabs (Hologic® Aptima 2 Combo, San Diego, CA) for extragenital (or vaginal, if applicable) GC/CT testing and two Tasso+™ devices. Mailing and tracking of kits is managed through Tasso’s confidential kit delivery portal. Study staff enter participant information into the portal, at which point kits are assembled and mailed directly to the participant’s specified address. Tracking numbers allow staff to track location of packages, when they are received at the home, and when the participant arranges for pick-up to return specimens to the clinic. An automated text and/or email is then generated to notify participants that kits have been shipped and a reminder is sent 7 days after delivery if kits have not yet been returned.

##### Telehealth visits and interim follow-up

Participants in the HB-PrEP arm have two options for completing scheduled telehealth follow-ups: (1) via telephone call with DRIS or (2) via HIPAA (Health Portability & Accountability Act)-compliant Zoom conference, which already exists within the SHC electronic medical record. Additionally, participants can reach study staff through WelTel – a secure and health information-compliant, two-way message support program already in use by DRIS in our clinic for contacting PrEP patients(35). Scheduled check-in visits for both arms consist of routine PrEP adherence counseling, screening for new HIV/STI symptoms, and PrEP prescription renewal if indicated. Per SHC protocols, participants with symptoms compatible with acute HIV or any STI must be evaluated in-person in lieu of receiving a home sampling kit.

#### Specimen Processing and Laboratory Evaluations

Individuals randomized to HB-PrEP are mailed sampling kits to their preferred address and return self-collected samples via prepaid express postal mail to the PHSKC laboratory for processing within 48 hours. Participants are advised to refrigerate any sample that cannot be shipped and received in clinic within 2 days (e.g., over weekend days). Upon receipt in the clinic, the study staff and PI conduct a brief quality assessment of specimens (i.e., ensure Aptima tubes are not open or leaking, etc.), place laboratory orders in the electronic health record, and send samples to the PHSKC laboratory for testing per standard clinic protocol. Additional processing and testing procedures for HIV, syphilis, RPR and creatinine have been described elsewhere.(30)

#### Outcomes and Analyses

##### Primary outcome measures

Effectiveness will be evaluated with a survival analysis using PrEP non-retention as the event of interest. Observation time begins for all participants at the time of randomization and continues until either loss-to-follow-up or study end at 20 months. Consistent with previous studies,(36,37) we define participants in either arm as “retained” on PrEP if they complete monitoring tests and visits with a clinician or DRIS at each 4-month interval to receive a renewed PrEP prescription. Kaplan-Meier curves will compare PrEP retention rates between the intervention and control arms at 20 months. We will use Cox proportional-hazards regression models to examine individual-level correlates (age, race/ethnicity, income range, education status, etc.) of PrEP retention.

##### Secondary outcome measures

1. Reach is defined as the proportion of individuals who choose to utilize HB-PrEP for at least half of tri-annual PrEP visits over the study period. As we are interested in the real-world uptake of HB-PrEP, we are tracking the number and proportion of individuals assigned to the intervention arm who choose at any time to transition to the SOC arm.
2. We adapted elements of the PPE-15,(38) an internationally validated questionnaire of patient experience and satisfaction, and questionnaires from other PrEP use trials for our tri-annual online participant surveys.
3. Study surveys inquire about PrEP use strategy (event-driven or “2-1-1” vs daily) over the preceding 4 months and adherence questions are tailored to the reported use strategy. Based on data that taking ≥4 doses/week confers a 96% reduction in HIV risk,(39) self-reported PrEP adherence is measured using validated question items asking the percent of PrEP taken in the prior 4 weeks.(40) The percentage of PrEP adherence will be compared between arms and analyzed using linear mixed models to account for repeated measures on each participant.
4. We will measure the time from receipt of monitoring and STI results into the SHC electronic health record to the time study staff notify the participant of abnormal results.
5. We will also track new syphilis and GC/CT infections for each group per tri-annual interval to estimate a composite asymptomatic STI positivity incident rate.

##### Qualitative data

Semi-structured interviews will be conducted with participants and study staff beginning at month 4 and occur at tri-annual intervals thereafter until a total of 24 participants have been interviewed. Purposive sampling will be employed to include participants who chose not to use HB-PrEP and groups with historically lower PrEP retention rates, such as Black MSM and younger individuals.(37) Three DRIS and administrators will be interviewed at 12 and 20 months to understand operational barriers and facilitators B&F of program implementation. Transcripts will be independently reviewed by two study staff and evaluated using thematic coding and a mix of inductive and deductive coding. Participants from the intervention arm will be invited to participate in interviews focused on understanding specific barriers and facilitators of PrEP retention, identifying unanticipated negative effects or issues with delivery of the intervention, determining whether HB-PrEP is preferred to SOC, and how future iterations of the program might be improved. Participation in interviews will be incentivized.

## Results

Enrollment for the trial began in March 2022 and continued through May 2022 when all research was halted in the SHC to focus on the emerging mpox outbreak. Study enrollment then resumed in December 2022 and is ongoing.

A total of 161 PrEP patients were screened to enroll the first 100 participants into the intervention arm (49%) or standard care arm (51%). Two PrEP patients (1.2%) were deemed ineligible to enroll, 55 (34%) declined to participate, and staff were not able to discuss enrollment with four (2.5%) patients. Common reasons for declining to participate included having a preference for coming into the clinic or not wanting to see or collect their own blood samples.

Table 2 describes demographics and other baseline characteristics participants self-reported upon enrollment. The baseline survey completion rate was 96% for both trial arms. Median age was 34 years (interquartile range: 28-39, overall range: 19-67). Of participants who elected to provide sex and gender information, nearly all were assigned male sex at birth (99%). Most identified as cisgender men (89%), gay (89%), and their race/ethnicity as non-Hispanic White (38%) or Asian (25%). The majority of participants reported an annual income of $30,000 or more (75%) and at least some college education (91%).

**Table 2:**
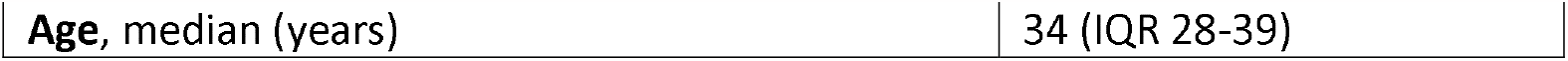
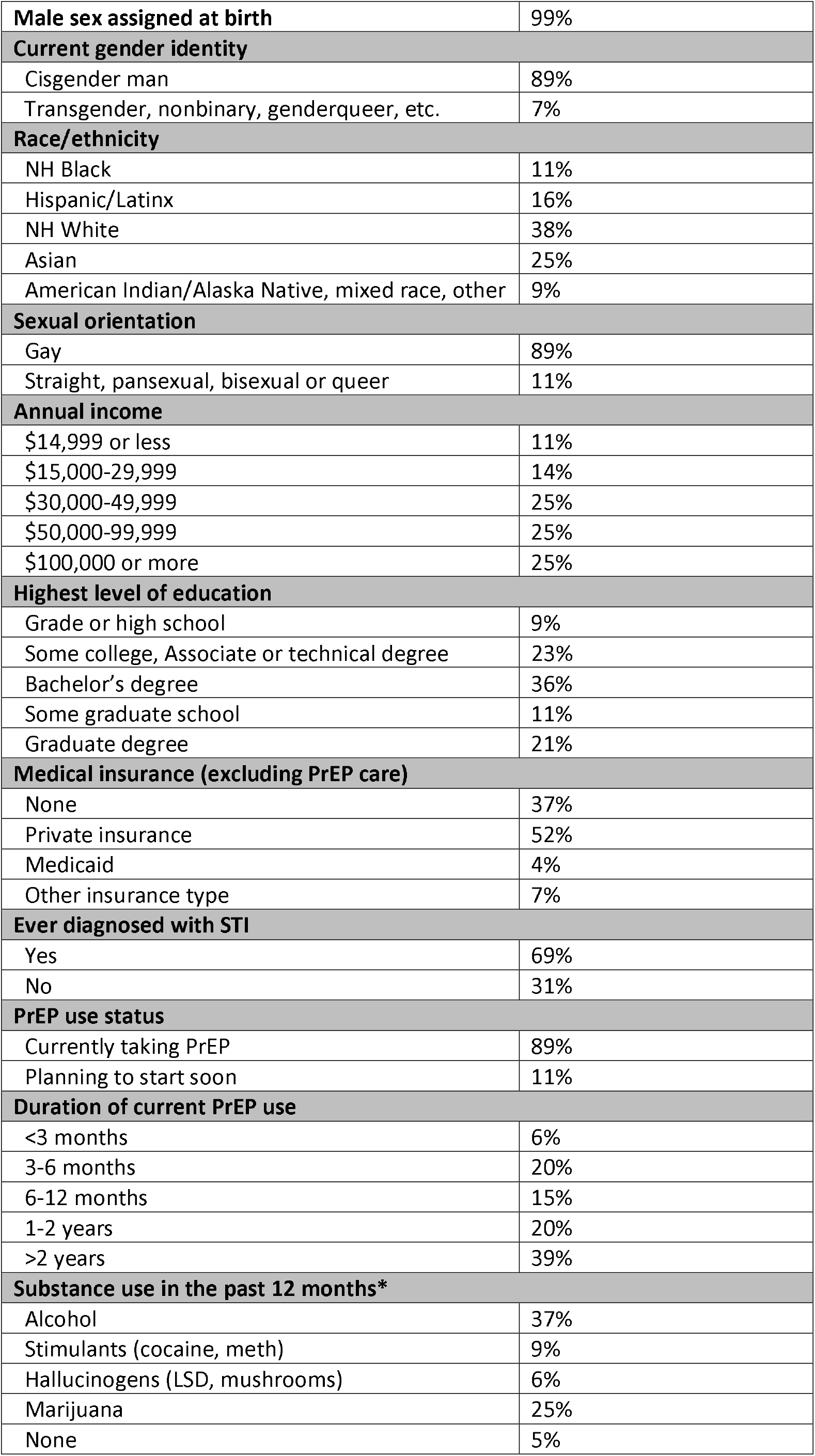
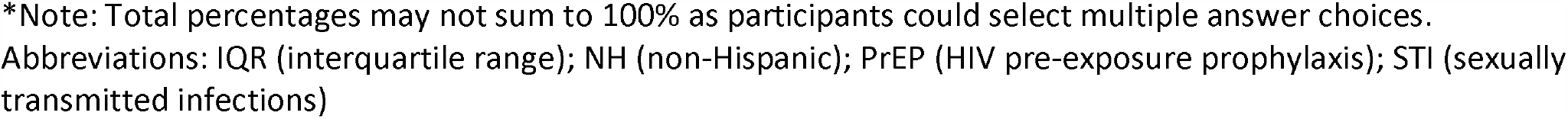
Baseline characteristics of 100 HOT4PrEP enrollees, March 2022 – May 2023.

|  |  |
| --- | --- |
| Age, median (years) | 34 (IQR 28-39) |
| **Male sex assigned at birth** | 99% |
| **Current gender identity** |  |
| Cisgender man | 89% |
| Transgender, nonbinary, genderqueer, etc. | 7% |
| **Race/ethnicity** |  |
| NH Black | 11% |
| Hispanic/Latinx | 16% |
| NH White | 38% |
| Asian | 25% |
| American Indian/Alaska Native, mixed race, other | 9% |
| **Sexual orientation** |  |
| Gay | 89% |
| Straight, pansexual, bisexual or queer | 11% |
| **Annual income** |  |
| \$14,999 or less | 11% |
| \$15,000-29,999 | 14% |
| \$30,000-49,999 | 25% |
| \$50,000-99,999 | 25% |
| \$100,000 or more | 25% |
| **Highest level of education** |  |
| Grade or high school | 9% |
| Some college, Associate or technical degree | 23% |
| Bachelor's degree | 36% |
| Some graduate school | 11% |
| Graduate degree | 21% |
| **Medical insurance (excluding PrEP care)** |  |
| None | 37% |
| Private insurance | 52% |
| Medicaid | 4% |
| Other insurance type | 7% |
| **Ever diagnosed with STI** |  |
| Yes | 69% |
| No | 31% |
| **PrEP use status** |  |
| Currently taking PrEP | 89% |
| Planning to start soon | 11% |
| **Duration of current PrEP use** |  |
| <3 months | 6% |
| 3-6 months | 20% |
| 6-12 months | 15% |
| 1-2 years | 20% |
| >2 years | 39% |
| **Substance use in the past 12 months\*** |  |
| Alcohol | 37% |
| Stimulants (cocaine, meth) | 9% |
| Hallucinogens (LSD, mushrooms) | 6% |
| Marijuana | 25% |
| None | 5% |

\*Note: Total percentages may not sum to 100% as participants could select multiple answer choices.
Abbreviations: IQR (interquartile range); NH (non-Hispanic); PrEP (HIV pre-exposure prophylaxis); STI (sexually transmitted infections)

All participants in the intervention arm were contacted prior to their 4-month follow-up visit to ask if they would prefer to come into clinic or receive home kits. Of the 49 participants contacted, 32 (65%) opted to collect samples at home and follow up via telehealth; 27 (84%) participants returned test kits with samples for HIV/STI testing.

## Discussion

The HOT4PrEP study is a hybrid implementation study to evaluate whether having the option of HB-PrEP improves PrEP retention over time among persons on PrEP in the Seattle-King County SHC. Preliminary data from the first 100 participants suggest that HB-PrEP monitoring – a strategy involving self-collected GC/CT swabs, blood specimens obtained using the Tasso device, and telehealth follow-up – is acceptable to users and feasible to implement into a busy PrEP clinic. Approximately two-thirds of participants who were offered the option of HB-PrEP chose to use it, and of those who received kits, 84% successfully returned kits containing samples sufficient to complete their requisite PrEP monitoring labs. Enrollment to the randomized portion of the trial is ongoing and study staff are recruiting for the qualitative component of the study. Analyses of preliminary PrEP retention rates at 4 and 8 months and secondary outcomes are underway. While not every person may choose to use a home-based option for every PrEP follow-up visit, HB-PrEP holds promise as a strategy to decrease barriers for staying on PrEP, increase patient autonomy and convenience, and expand capacity of clinics to initiate and maintain a greater number of people on PrEP.

## Data Availability

All preliminary data produced in the present work are contained in the manuscript.

